# Seroresponse to SARS-CoV-2 vaccines among maintenance dialysis patients

**DOI:** 10.1101/2021.08.19.21262292

**Authors:** Caroline M. Hsu, Daniel E. Weiner, Gideon N. Aweh, Harold J. Manley, Vladimir Ladik, Jill Frament, Dana Miskulin, Christos Agyropoulos, Kenneth Abreo, Andrew Chin, Reginald Gladish, Loay Salman, Doug Johnson, Eduardo K. Lacson

## Abstract

**Importance:** Vaccines against SARS-CoV-2 are highly effective in the general population; however, their efficacy may be diminished in maintenance dialysis patients, a population particularly vulnerable to COVID-19 infection and morbidity.

**Objective:** We assessed vaccine response in a national sample of maintenance dialysis patients and identified predictors of response.

**Design:** Retrospective cohort study

**Setting:** 130 Dialysis Clinic, Inc (DCI) facilities

**Participants:** Maintenance dialysis patients without known prior COVID-19 or a positive baseline antibody titer

**Exposure(s):** Vaccine type and clinical characteristics

**Main Outcome(s):** Using a semi-quantitative assay for antibodies against SARS-CoV-2 spike antigen, vaccine response was defined as at least one titer ≥1 U/L between 14 and 74 days after completion of a SARS-CoV-2 vaccine series. Regression analysis was used to identify characteristics associated with response.

**Results:** Among 1528 patients, 437 received BNT162b2/Pfizer vaccine, 766 received mRNA-1273/Moderna, and 325 received Ad26.COV2.S/Janssen. Serologic response differed significantly by vaccine type: 381/437 (87%) among BNT162b2/Pfizer recipients, 736/766 (96%) among mRNA-1273/Moderna recipients, and 119/325 (37%) among Ad26.COV2.S/Janssen recipients. Vaccine type, older age, immune-modulating medication, history of transplantation, and lower serum albumin were associated with vaccine non-response.

**Conclusions and Relevance:** Serologic response to mRNA vaccines is robust among maintenance dialysis patients. Future research should evaluate durability of this response, correlation between seroresponse and protection from COVID-19, and the role of the AD26.COV2.S/Janssen vaccine in this vulnerable population.

## Introduction

As of August 2021, three vaccines against SARS-CoV-2 have received Emergency Use Authorization (EUA) by the United States Food and Drug Administration (FDA): two mRNA vaccines (BNT162b2/Pfizer and mRNA-1273/Moderna) and one adenoviral vector vaccine (Ad26.COV2.S/Janssen). All three vaccines are highly effective in the general population,^1–3^ and their widespread deployment has shifted the course of the COVID-19 pandemic.

Studies to date suggest a high seroresponse rate (greater than 80%) to mRNA vaccines in maintenance dialysis patients, albeit less than that in the general population.^4–7^ A report in 76 dialysis patients that Ad26.COV2S elicits lower seroresponse warrants further study;^8^ moreover, small sample sizes have so far limited evaluation of predictors of non-response. Accordingly, we retrospectively analyzed seroresponse to SARS-CoV-2 vaccines in maintenance dialysis patients, updating an earlier report.^9^

## Methods

Dialysis Clinic, Inc. (DCI) is a national not-for-profit provider that cares for more than 15,000 patients at 260 outpatient dialysis clinics across 29 states. Beginning in January 2021, DCI physicians had the option of activating a SARS-CoV-2 vaccine protocol, modeled after the existing hepatitis B vaccine protocol whereby antibody titers are measured to document seroresponse to vaccination. Treating physicians could activate the protocol upon patient receipt of SARS-CoV-2 vaccine, regardless of vaccine type or site of administration. As part of the monthly blood draws beginning after receipt of a SARS-CoV-2 vaccine, immunoglobulin G spike antibodies (SAb-IgG) against the receptor-binding domain of the S1 subunit of SARS-CoV-2 spike antigen were measured using the chemiluminescent assay ADVIA Centaur® XP/XPT COV2G, which received emergency use authorization in July 2020.^10^ This semi-quantitative assay has a range between 0 and ≥20 U/L, and SAb-IgG titer ≥1 U/L signifies patient seroresponse.^10^

Demographic and clinical data, vaccination dates, and SAb-IgG titer results were obtained from the DCI electronic health record. Patients with previously diagnosed COVID-19 were excluded, as were patients with SAb-IgG titer ≥1 U/L before vaccination or within 10 days after vaccination for likely prior undiagnosed COVID-19.

In primary analyses, we determined the SARS-CoV-2 vaccine seroresponse rates at 14 to 74 days after completion of a vaccine series, as well as time to peak titer within this time frame. This timing of SAb-IgG titer assessment (at least 14 days after completion of vaccine series) corresponds with the currently accepted definition of “fully vaccinated.”^11^ Multivariable log Poisson regression with robust variances was used to analyze the association of demographic and clinical factors with vaccine seroresponse. Secondary analyses used alternate definitions of vaccine seroresponse: (1) utilizing SAb-IgG ≥2 U/L (a threshold suggested by DCI’s internal validation methods), and (2) utilizing the first SAb-IgG titer measured at least 14 days after completion of a vaccine series. This study was reviewed and approved by the WCG IRB Work Order 1-1456342-1. Statistical analyses were performed using SAS v9.4.

## Results

Between February 1 and July 6, 2021, 1528 patients (437 [29%] BNT162b2/Pfizer recipients, 766 [50%] mRNA-1273/Moderna recipients, and 325 [21%] Ad26.COV2.S/Janssen recipients) across 130 dialysis facilities had SAb-IgG titers measured after receiving SARS-CoV-2 vaccination. 1029 (67%) patients had more than one titer checked between 14 and 74 days after completion of the vaccine series. Patients who received BNT162b2/Pfizer tended to be older, likely reflecting its earlier EUA and rollout. (**Table**) Vaccine seroresponse varied significantly by vaccine received: 381/437 (87%) of BNT162b2/Pfizer recipients, 736/766 (96%) of mRNA-1273/Moderna recipients, and 119/325 (37%) of Ad26.COV2.S/Janssen recipients. Median [interquartile range] number of days to peak titer from completion of the vaccine series was 33 [20-42], 29 [19-40], and 47 [34-61] days for the 3 vaccines, respectively. At titer ≥2 U/L, response rate was slightly lower for all vaccine types, with the greatest difference occurring in Ad26.COV2.S/Janssen recipients. When utilizing first SAb-IgG titer at least 14 days after completion of a vaccine series, seroresponse rate was significantly lower only among Ad26.COV2.S/Janssen recipients. (**Supplemental Table**).

**Table.** Patient characteristics by vaccine administered.

**Table. Patient characteristics by vaccine administered**
| Demographics | All Patients<br>(N=1,528) | BNT162b2/Pfizer<br>(N=437) | mRNA-1273/Moderna<br>(N=766) | Ad26.COV2.S/Janssen<br>(N=325) | P-value |
| --- | --- | --- | --- | --- | --- |
| Age (years) | 64.2 ± 13.5 | 66.8 ± 12.9 | 64.0 ± 13.9 | 61.3 ± 12.6 | <0.001 |
| Female | 646(42.3) | 191(43.7) | 308(40.2) | 147(45.2) | 0.24 |
| Race |  |  |  |  | <0.001 |
| White | 768(50.3) | 198(45.3) | 447(58.4) | 123(37.8) |  |
| Black | 356(23.3) | 80(18.3) | 116(15.1) | 160(49.2) |  |
| Native American | 126(8.3) | 61(14.0) | 60(7.8) | 5(1.5) |  |
| Asian/Pacific Islander | 93(6.1) | 49(11.2) | 37(4.8) | 7(2.2) |  |
| Other/Unknown | 185(12.1) | 49(11.2) | 106(13.8) | 30(9.2) |  |
| Hispanic | 205(13.4) | 63(14.4) | 134(17.5) | 8(2.5) | <0.001 |
| Vintage (months) | 53.0 ± 58.1 | 50.9 ± 57.6 | 52.4 ± 57.0 | 57.4 ± 61.2 | 0.28 |
| Body Mass Index (kg/m<sup>2</sup>) | 28.4 ± 7.1 | 28.4 ± 7.4 | 28.1 ± 6.4 | 29.2 ± 8.0 | 0.06 |
| Long Term Care Facility | 139(9.1) | 41(9.4) | 69(9.0) | 29(8.9) | 0.97 |
| Home Dialysis | 224(14.7) | 59(13.5) | 130(17.0) | 35(10.8) | 0.02 |
| Peritoneal Dialysis | 198(13.0) | 50(11.4) | 113(14.8) | 35(10.8) |  |
| Home Hemodialysis | 26(1.7) | 9(2.1) | 17(2.2) | 0(0.0) |  |
| Adequate Dialysis Dose<sup>a</sup> | 1,205(78.9) | 350(80.1) | 597(77.9) | 258(79.4) | 0.66 |
| Serum Albumin (g/dl) | 3.9 ± 0.4 | 3.9 ± 0.4 | 3.9 ± 0.4 | 3.8 ± 0.4 | 0.05 |
| Other Vaccines within 14 days | 129(8.4) | 40(9.2) | 62(8.1) | 27(8.3) | 0.81 |
| Pneumococcal | 32(2.1) | 5(1.1) | 21(2.7) | 6(1.9) | 0.18 |
| Hepatitis B | 101(6.6) | 35(8.0) | 44(5.7) | 22(6.8) | 0.31 |
| Flu Vaccine | 6(0.4) | 2(0.5) | 4(0.5) | 0(0.0) | 0.99 |
| Hepatitis B seroimmunity<sup>b</sup> | 1,099(71.9) | 317(72.5) | 557(72.7) | 225(69.2) | 0.34 |
| Potential Immunosuppression | 276(18.1) | 94(21.5) | 126(16.5) | 56(17.2) | 0.08 |
| Immune-modulating Medications | 197(12.9) | 69(15.8) | 91(11.9) | 37(11.4) | 0.10 |
| Prior Transplant | 102(6.7) | 32(7.3) | 53(6.9) | 17(5.2) | 0.49 |
| Immunodeficiency Disorder | 71(4.7) | 23(5.3) | 31(4.1) | 17(5.2) | 0.54 |
| Hospitalization within 14 days | 194(12.7) | 58(13.3) | 103(13.5) | 33(10.2) | 0.30 |
| Disability | 62(4.1) | 16(3.7) | 30(3.9) | 16(4.9) | 0.66 |
| Tobacco Use | 205(13.4) | 48(11.0) | 101(13.2) | 56(17.2) | 0.04 |
| Alcohol Abuse Disorder | 143(9.4) | 38(8.7) | 71(9.3) | 34(10.5) | 0.71 |
| Drug Abuse Disorder | 51(3.3) | 14(3.2) | 27(3.5) | 10(3.1) | 0.92 |
| Number of Comorbidities | 2.9 ± 1.7 | 2.9 ± 1.6 | 2.9 ± 1.6 | 3.1 ± 1.8 | 0.10 |
| Diabetes Mellitus | 909(59.5) | 269(61.6) | 437(57.1) | 203(62.5) | 0.15 |
| Hypertension | 1,310(85.7) | 371(84.9) | 657(85.8) | 282(86.8) | 0.77 |
| Congestive Heart Failure | 298(19.5) | 72(16.5) | 150(19.6) | 76(23.4) | 0.06 |
| COPD<sup>c</sup> | 230(15.1) | 65(14.9) | 100(13.1) | 65(20.0) | 0.01 |
| Stroke/Cerebrovascular Disorder | 114(7.5) | 33(7.6) | 51(6.7) | 30(9.2) | 0.34 |
| Peripheral Vascular Disease | 178(11.7) | 43(9.8) | 78(10.2) | 57(17.5) | 0.001 |
| Thyroid Disorder | 248(16.2) | 64(14.7) | 136(17.8) | 48(14.8) | 0.27 |
| History of Cancer | 136(8.9) | 41(9.4) | 74(9.7) | 21(6.5) | 0.22 |

<sup>a</sup> Adequate dialysis defined by hemodialysis single pool Kt/V≥1.2 or peritoneal dialysis weekly Kt/V≥1.7.
<sup>b</sup> Hepatitis B seroimmunity defined as hepatitis B surface antibody ≥ 10 mIU/mL
<sup>c</sup> COPD Chronic Obstructive Pulmonary Disease
Hepatitis B seroimmunity defined as hepatitis B surface antibody $\geq 10$ mIU/mL

In multivariable analysis, vaccine type, older age, non-Black and non-Native American race, immune-modulating medications, history of transplantation, and lower serum albumin were associated with lower likelihood of vaccine seroresponse (**Figure**).

**Figure.**
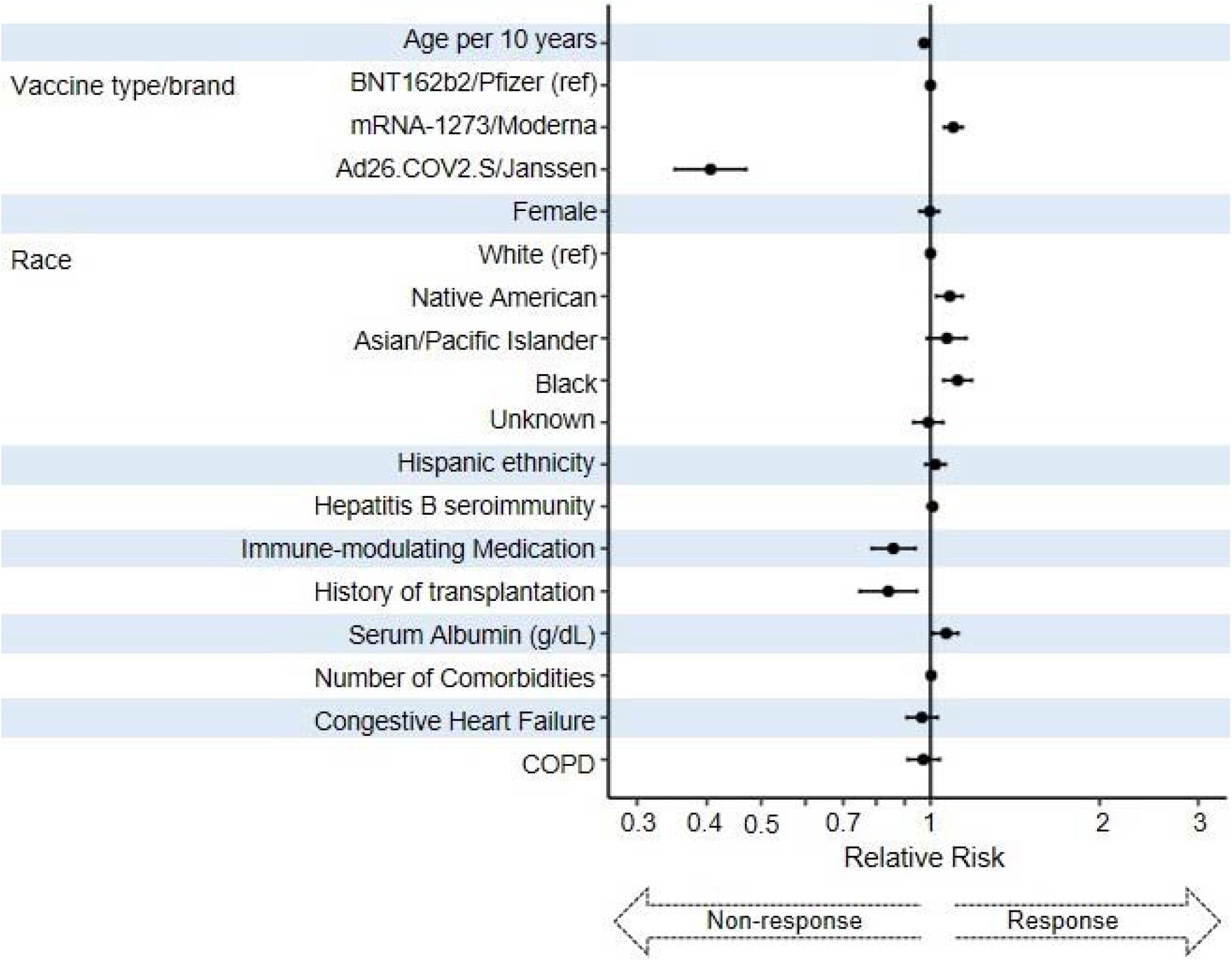
Multivariable regression of clinical characteristics predicting SARS-CoV-2 vaccine response. SARS-CoV-2 vaccine response defined as immunoglobulin-G spike antibodies (SAb-IgG) against the receptor-binding domain of the S1 subunit of SARS-CoV-2 spike antigen titer ≥1 U/L measured 14-74 days after completion of a vaccine series Hepatitis B seroimmunity defined as hepatitis B surface antibody ≥10 mIU/mL COPD Chronic Obstructive Pulmonary Disease Analysis was performed with multivariable log Poisson regression with robust variances.

## Discussion

Among maintenance dialysis patients, mRNA vaccines against SARS-CoV-2 elicited appropriate seroresponse in the vast majority, consistent with prior reports of mRNA vaccines worldwide.^4–7^ In contrast, seroresponse to the Ad26.COV2.S/Janssen vaccine was low, consistent with an earlier small study,^8^ and suggesting that mRNA-based SARS-CoV-2 vaccines should be recommended for maintenance dialysis patients, particularly given their high morbidity and mortality from COVID-19.^12^

The comparatively low seroresponse rate to the Ad26.COV2.S/Janssen vaccine among the dialysis population is concerning. SAb-IgG antibodies are thought to confer protection from COVID-19 itself via neutralization of the spike protein, and post-vaccination antibody titers have been shown to correlate with protection from COVID-19.^13^ As an adenoviral vector vaccine, the Ad26.COV2.S/Janssen vaccine must enter the cell nucleus and undergo transcription, an extra step compared to mRNA vaccines. Of note, the timing of SAb-IgG titer assessment may have affected results; as a single-dose regimen, the response to Ad26.COV2.S here is assessed 3-4 weeks earlier relative to the initial vaccine dose than is the response to mRNA vaccines. The greater seroresponse by titer within 74 days versus by first titer suggests that the Ad26.COV2.S/Janssen vaccine’s seroresponse may continue to increase beyond two weeks post-vaccination. However, long-term data from the phase 1-2a trial indicate that titers eventually level off and even decrease slightly.^14^ Thus, even allowing for later increase in seroresponse, the Ad25.COV2.SS/Janssen vaccine appears to have significantly lower efficacy than the mRNA vaccines among maintenance dialysis patients.

While only Ad26.COV2.S/Janssen’s single low-dose regimen currently has emergency use authorization in the United States, the phase 1-2a trial had included two-dose regimens and high-dose regimens which elicited higher titers, albeit with more adverse events.^15^ Whether a second dose may provide benefit to certain populations remains under investigation (ClinicalTrials.gov number NCT04614948). The difference between BNT162b2/Pfizer and mRNA-1273/Moderna was less extreme but still statistically significant and may be related to differences of dosage (100 μg vs 30 μg of mRNA content, respectively), or, given the earlier availability of the BNT162b2/Pfizer vaccine, unaccounted-for confounding factors. Admittedly, the SAb-IgG antibody titer needed for protection from COVID-19 and the role of vaccine-induced cellular immunity remain uncertain, issues that are also complicated by emerging variants. Other than vaccine type, predictors of vaccine non-response were largely factors related to potential immunocompromise, including increasing age, the presence of immune-modulating medication, and lower serum albumin, all of which have been noted by others.^4–6^

Our study has its limitations. In this observational retrospective study, some confounders may not have been accounted for. The comparison of maximum to initial SAb-IgG titers is limited by the patients who only had one titer checked between 14 and 74 days. Lastly, as noted, while the assessment of SAb-IgG titer at least 14 days after completion of vaccine series corresponds with the current definition of “fully vaccinated,”^11^ it creates a difference in timing relative to the initial dose between one-dose and two-dose regimens.

In conclusion, mRNA vaccines are associated with greater seroresponse in maintenance dialysis patients, and they should continue to be widely used to protect this vulnerable population. Further study is needed to monitor the durability of this seroresponse. In addition, further investigation should evaluate the role of the Ad26.COV2.S/Janssen vaccine in this population, and repeat dosing may be needed.

## Data Availability

These data are not available for sharing

