## Supplemental Table for "Seroresponse to SARS-CoV-2 vaccines among maintenance dialysis patients"

**Supplemental Table**. Response rates by vaccine type, with sensitivity analyses for different definitions of seroresponse

|  | Definitions of seroresponse | | |
| --- | --- | --- | --- |
|  | Titer ≥1 U/L  14-74 days after completion of vaccine series | Titer ≥2 U/L  14-74 days after completion of vaccine series | Titer ≥1 U/L  at first measurement at least 14 days after completion of vaccine series |
| BNT162b2/Pfizer | 381/437 (87%) | 361/437 (83%) | 380/437 (87%) |
| mRNA-1273/Moderna | 736/766 (96%) | 720/766 (94%) | 736/766 (96%) |
| Ad26.COV2.S/Janssen | 119/325 (37%) | 85/325 (26%) | 83/325 (26%) |

Titer refers to the level of immunoglobulin-G spike antibodies (SAb-IgG) against the receptor-binding domain of the S1 subunit of SARS-CoV-2 spike antigen
